# Brief report: Determinants of potential sexual activity reduction in the face of the Monkeypox epidemic

**DOI:** 10.1101/2022.08.01.22278287

**Authors:** Haoyi Wang, Kennedy J.I. d’Abreu de Paulo, Thomas Gültzow, Hanne M.L. Zimmermann, Kai. J. Jonas

## Abstract

The current monkeypox epidemic is most prevalent among men-who-have-sex-with-men (MSM). Vaccination programs are being rolled-out to curb the epidemic. Behavioural measures have been called for as well, e.g., by the WHO to reduce the number of sexual partners and sexual activity. We investigated intentions and determinants among Dutch MSM to follow such measures. Early July 2022, 394 MSM answered an online questionnaire. The overall intentions to reduce number of partners and sexual activity was high, but only a minority had developed definite intentions. Determinant analysis revealed that dating/open relationship status was a positive predictor, vaccination intentions did not predict sexual behaviour change; those not on PrEP were more likely to change their sexual behaviour. Monkeypox infection concern was negatively related to weaker intentions and only predicted definite intentions. Our results show that additional public health measures are necessary to reach and convince MSM to engage in sexual behaviour change.

## Introduction

While monkeypox was known as a rare zoonotic disease caused by an orthopoxvirus leading to symptoms in humans similar to smallpox (1), it has recently changed from infections predominantly due to an interaction or activity with animals to human-to-human transmission (1, 2) triggering a global epidemic. At present day monkeypox is primarily affecting men-who-have-sex-with-men (MSM) in European countries and the number of infections are still increasing (3, 4). In the Netherlands 878 cases (as of 28.07.2022) have been reported, with the majority in Amsterdam (4).

To curb the spread of the epidemic, vaccination programs have commenced in many countries that are focussing on high-risk populations, such as MSM who are using HIV pre-exposure prophylaxis (PrEP) (5). Given vaccine scarcity (6), it is paramount to identify highest at-risk populations, but also to employ additional prevention measures. Due to the incubation period of 9 days in the case of invasive exposure (7), and a one-to-three days latency until the typical lesions appear, it is difficult to detect infections quickly. Thus, WHO has recently called upon MSM to reduce their number of sex partners and sexual encounters as an additional measure to prevent further spreading of infections (8, 9).

To our knowledge no findings regarding the willingness to reduce the number of sex partners and sexual activities among MSM and its determinants have been reported for the context of monkeypox. Yet, these results are relevant to inform the potential effect of the recommendation, and further national responses above and beyond current measures. Therefore, we aimed to investigate the willingness and determinants of sexual behaviour change in MSM to potentially reduce further monkeypox transmissions.

## Methods

### Participants and Procedure

To investigate our research question, we conducted an online survey among 394 MSM using a cohort established in 2017 (n=257), along with recruitment of MSM on a gay online dating app (n=137) in the first half of July 2022, prior to the start of structural monkeypox vaccination in the Netherlands (10). The study was assessed and approved by the ERCPN of Maastricht University (ref.188_11_02_2018_S32). Informed consent was provided by all participants.

### Measures

Participants were asked whether they would be willing to reduce 1) the number of sexual partners (hereinafter fewer partner intentions) and 2) the number of sexual encounters in the context of the monkeypox epidemic (hereinafter less sex intentions). These two endpoints were measured on a 1-5 Likert scale (with 1 = “definitely not” and 5 “definitely yes”). In this study, we categorised these two endpoints with two conceptualisations: a) “probably (4) or definitely (5) intending to reduce” vs. the rest of the scale points (1-3) and b) “definitely (5) intending to reduce” vs. the rest of the scale points (1-4).

We also assessed sociodemographic, behavioural and psycho-social determinants, which included: age, relationship status, education, employment status, migration status, place of residence, number of sex partners in the previous six months, HIV status, PrEP use status, substance use status in the previous six months, and gay subculture/sexual activities in the previous six months, knowing anybody who has/had monkeypox, monkeypox vaccination intentions, concern about being infected with monkeypox, perceived risk of being infected with monkeypox and perceived problematic consequences of monkeypox (see Table 1 and 2 for variables’ categorisations and measurements).

**Table 1.**
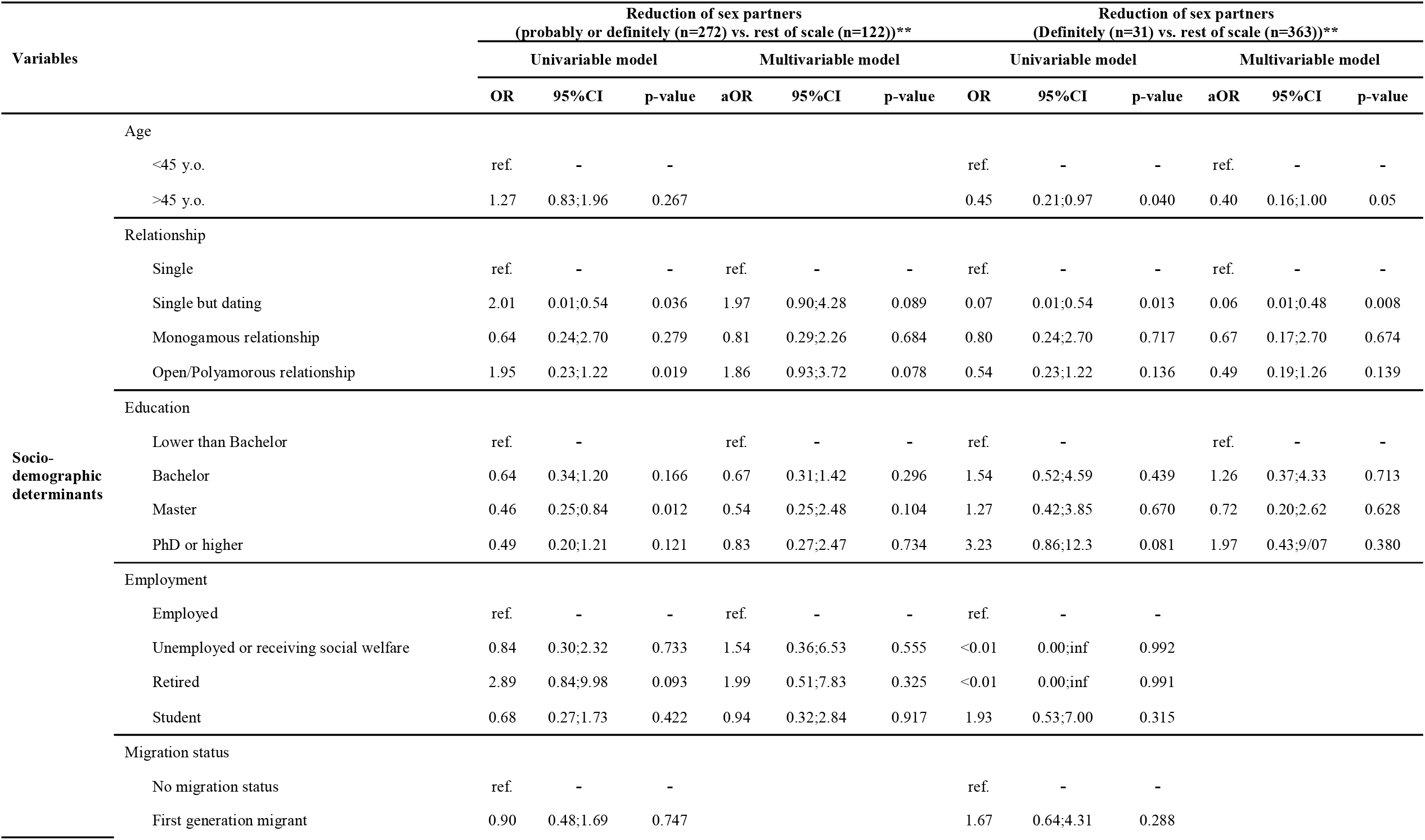
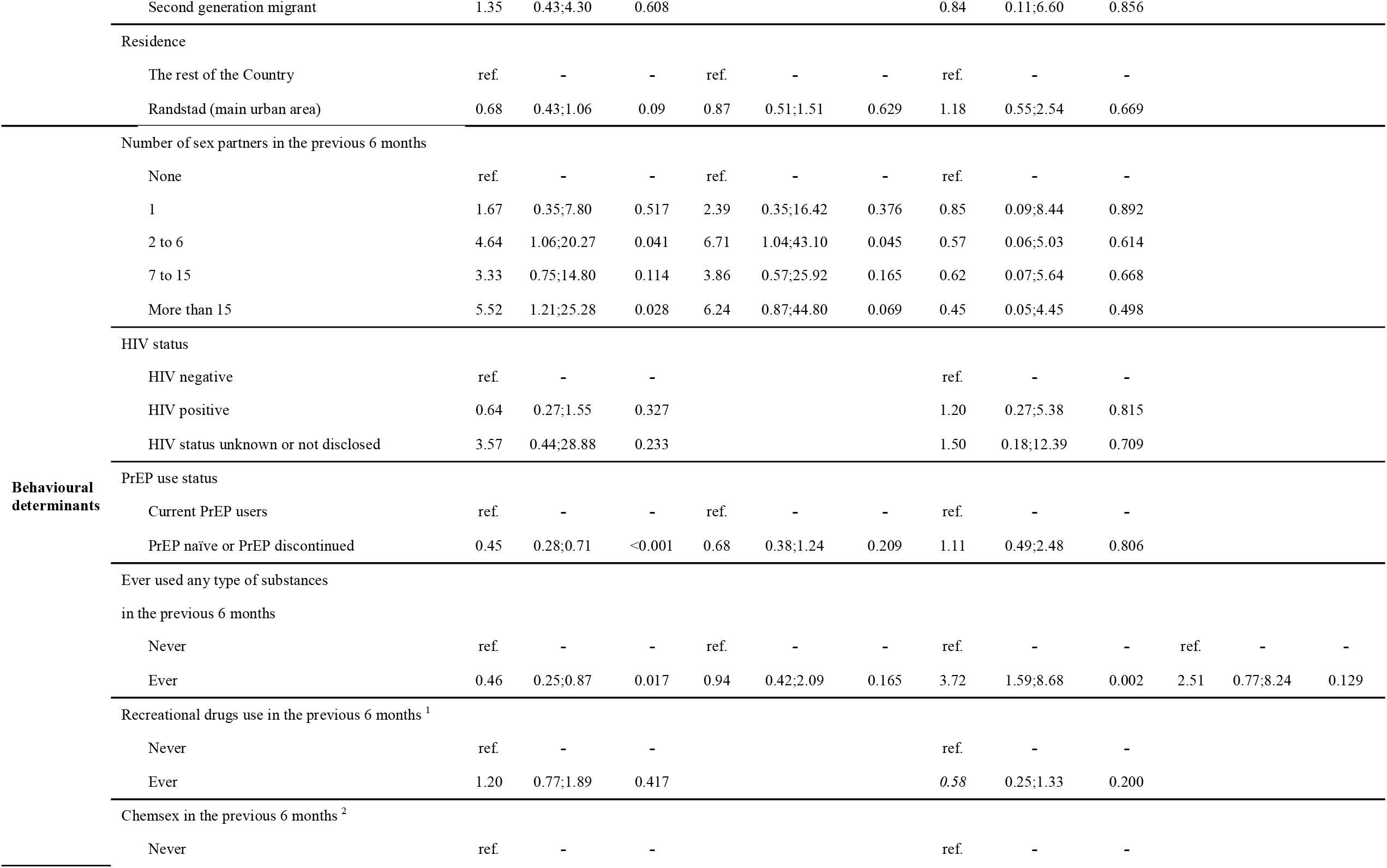
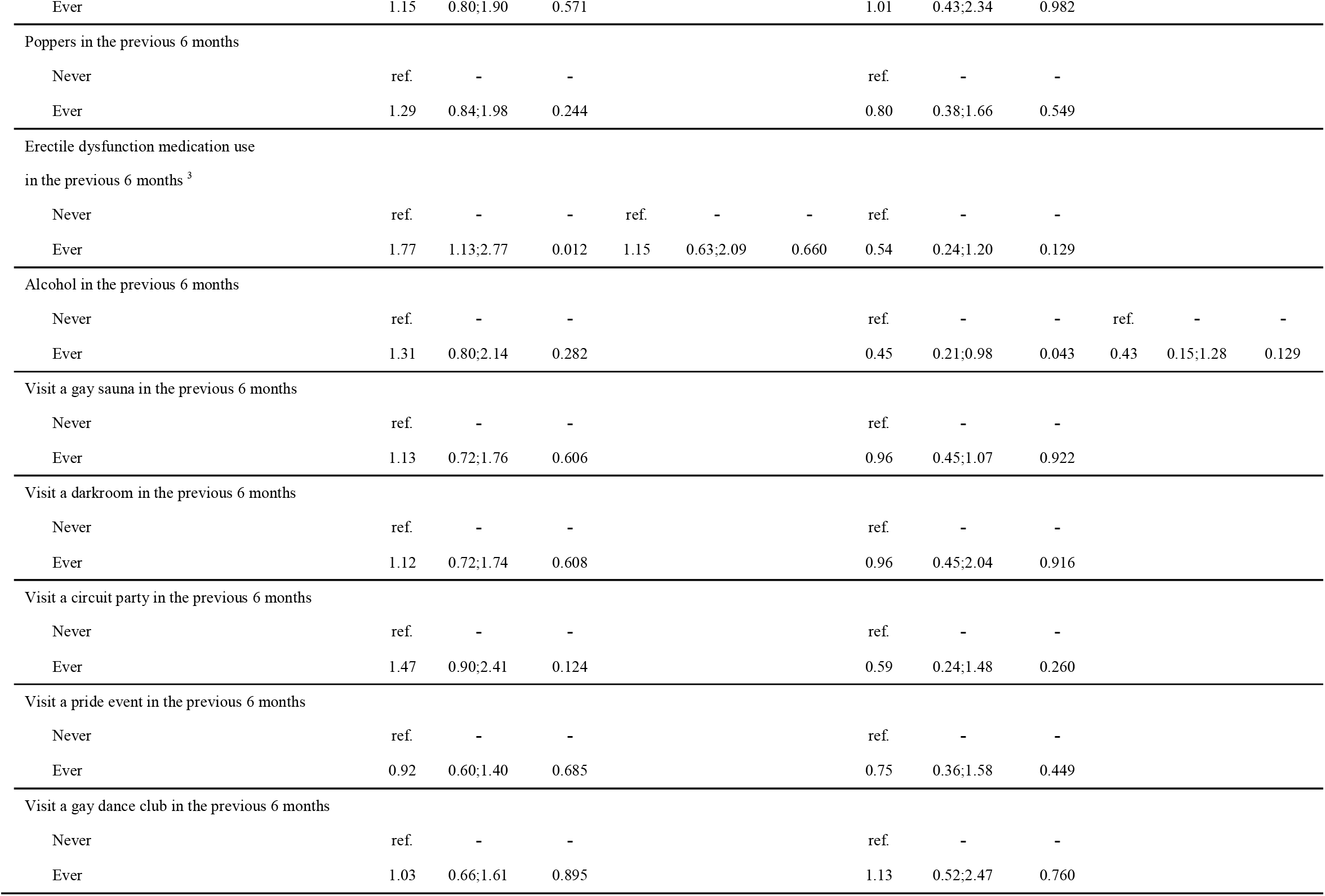
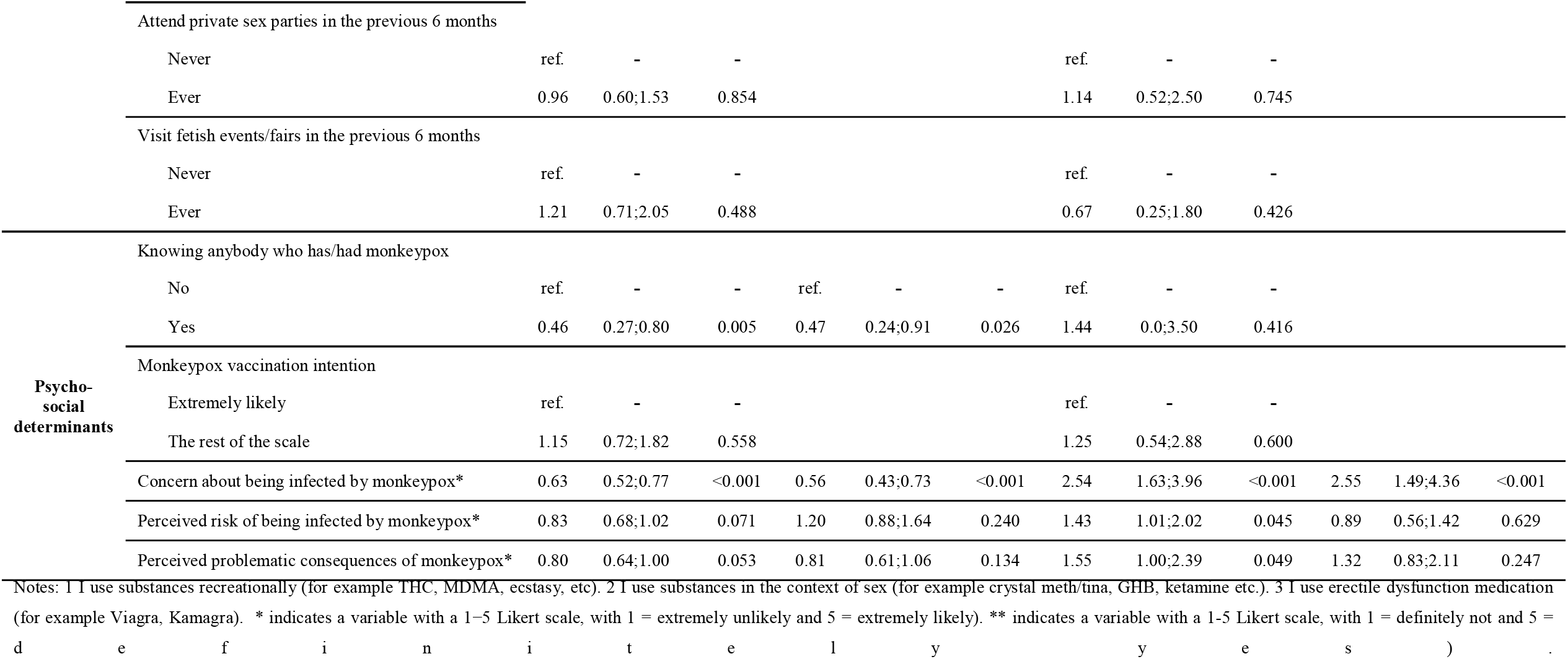
Determinants of intention to reduce the number of sex partners.

**Table 2.**
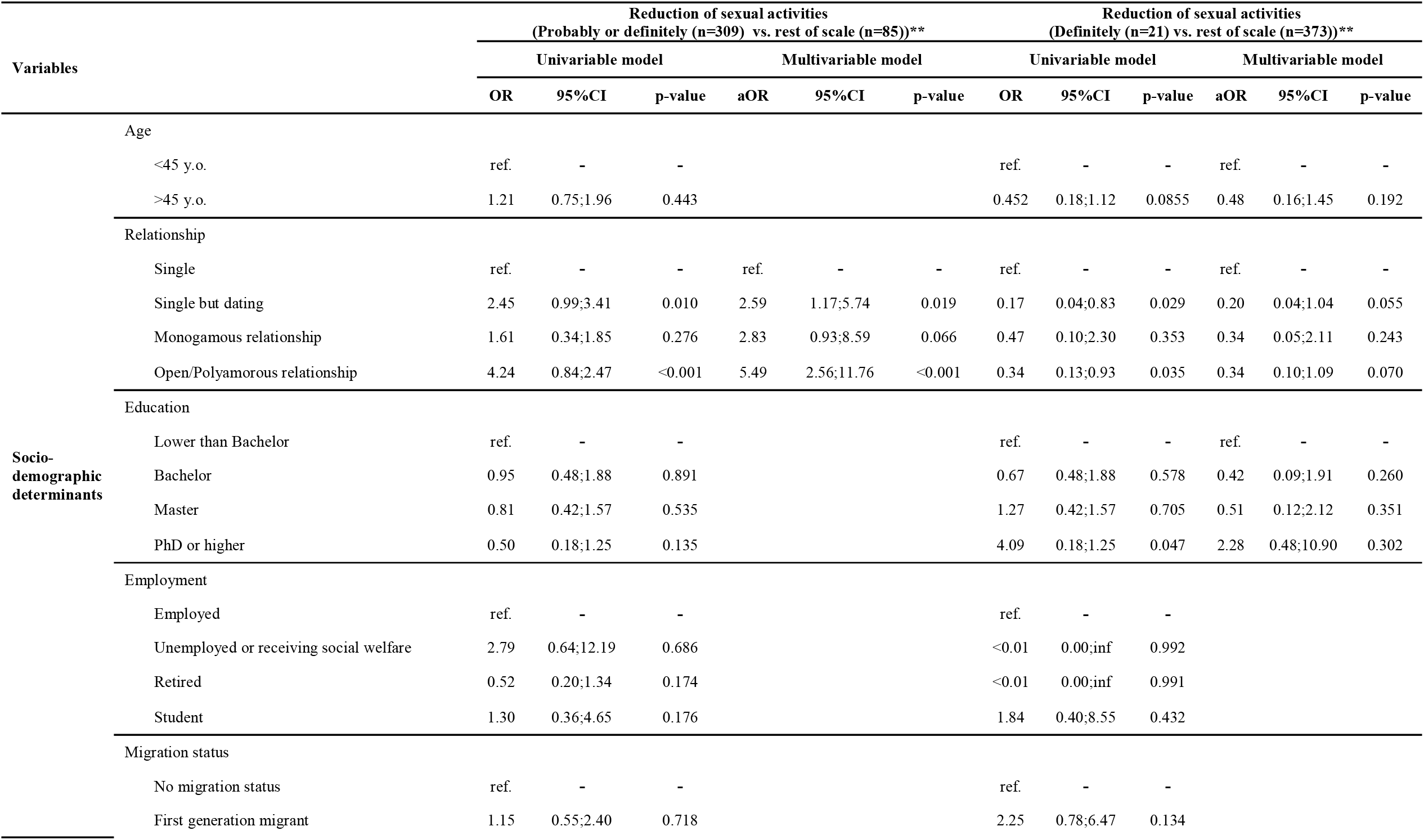
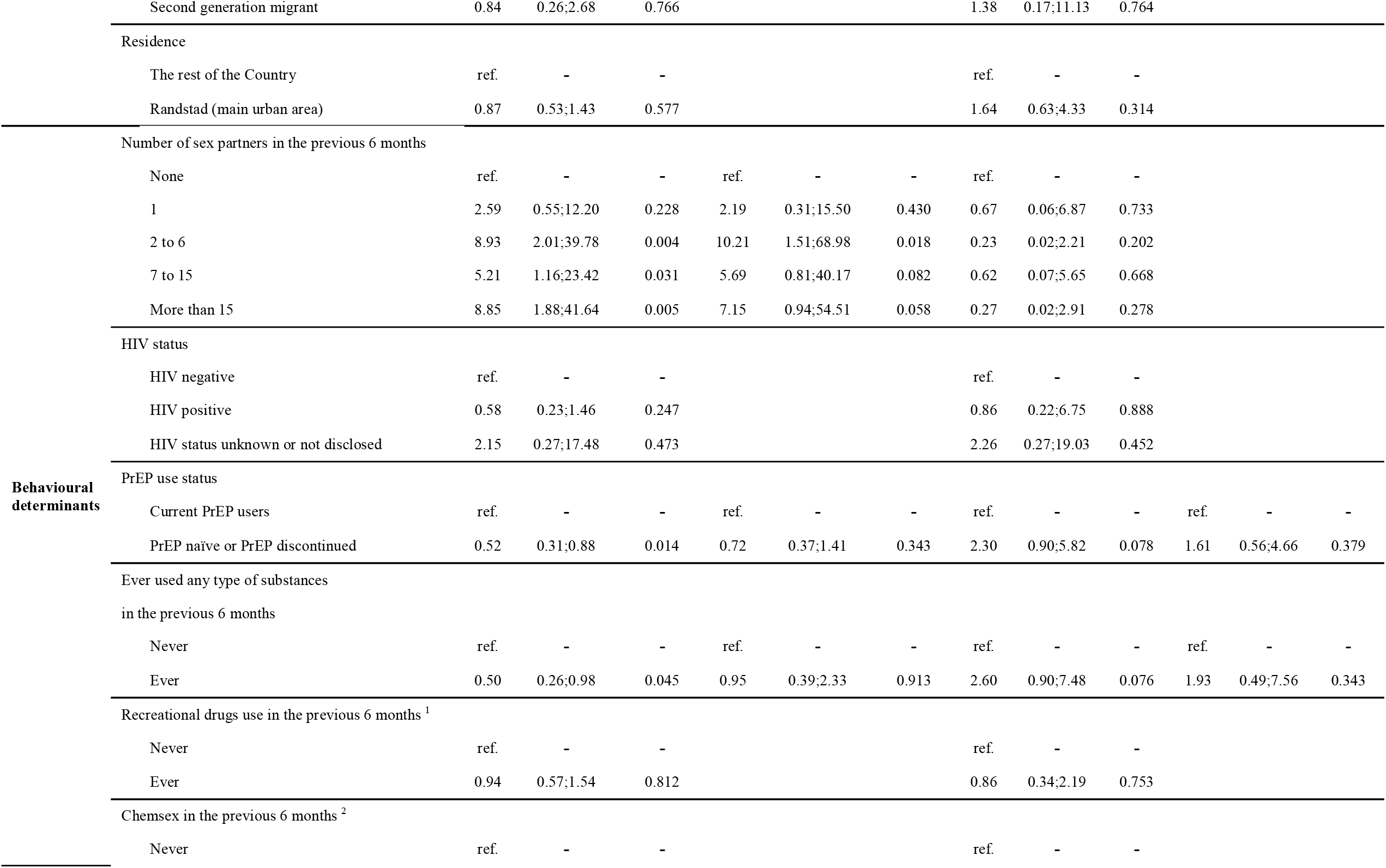
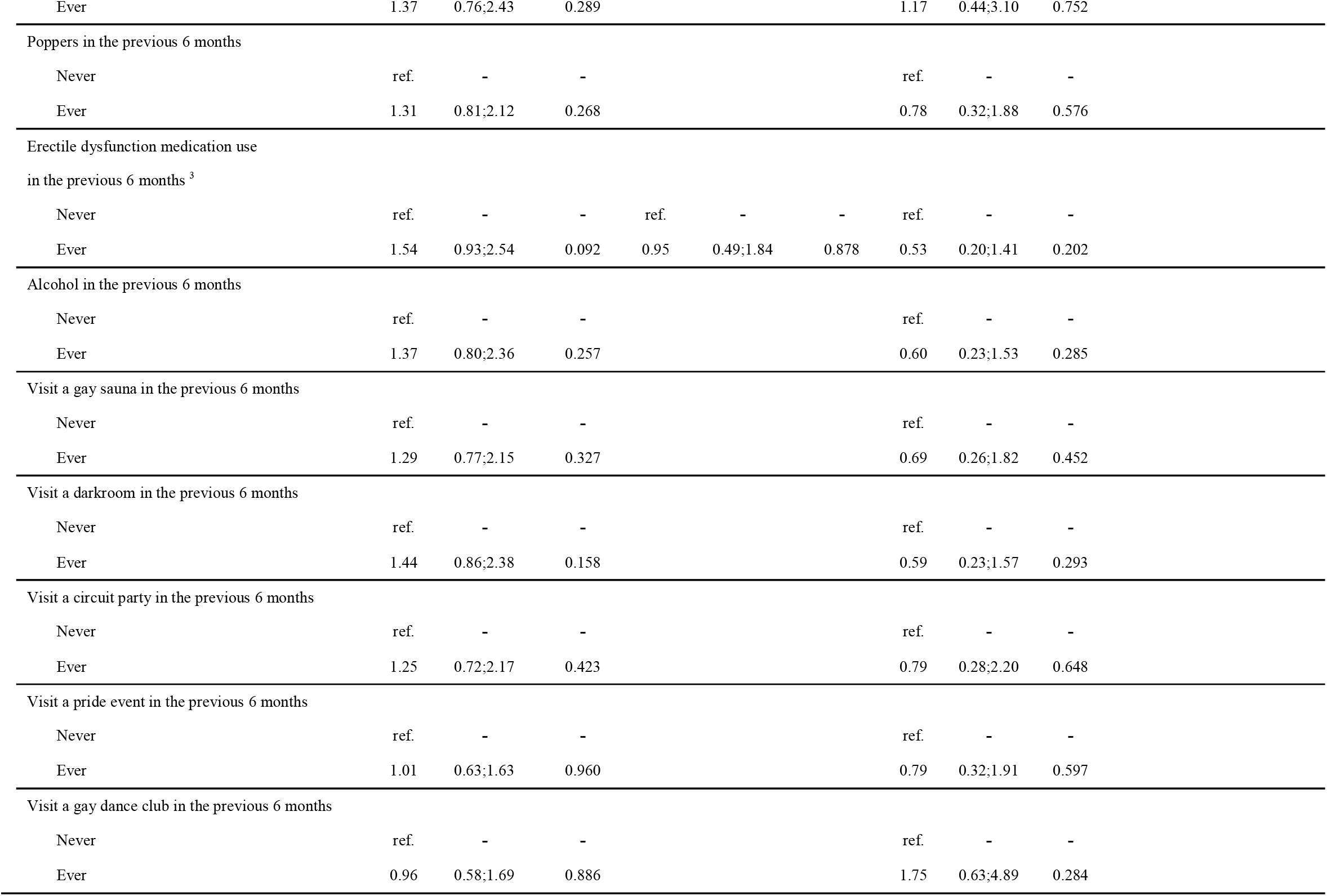
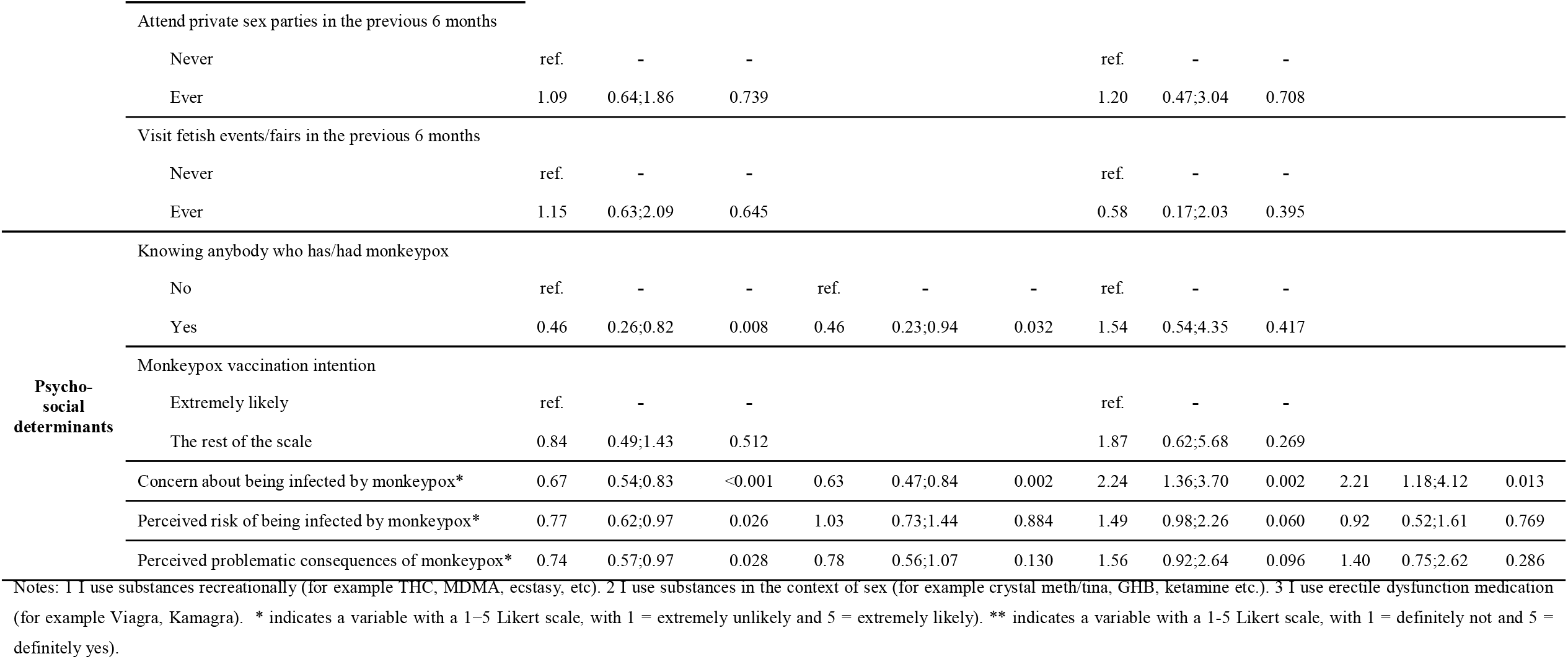
Determinants of intention to reduce the number of sexual activities.

### Statistical analysis

We conducted two multivariable logistic regression analyses with socio-demographic, behavioural, and psychosocial determinants for each endpoint. First, we conducted an univariable logistic model with each of the selected determinants to investigate potential correlations with the intention to have fewer partners/less sex given the monkeypox epidemic. All determinants with p<0.10 were retained in the multivariable model, given the relatively small sample size. Subsequently, we compared the results between different definitions of the endpoint to explore the implications on the endpoint selection to support future public health activities (see Table 1 and 2 for included determinants and models’ details). All analyses were conducted in R (version R 4.0.4).

## Results

### Study population characteristics

Of the included 394 MSM, 43% were below the age of 45, 6% were living with HIV and 66% were currently using PrEP (for the other population characteristics by PrEP use status see Wang et al. (11) as this study uses overlapping determinants, but different endpoints).

### Intention and determinants of the reduction of the number of sex partners

For fewer partner intentions as an endpoint, 272 (69%) MSM showed probable or definite intentions but only 31 (8%) MSM showed definite intentions to reduce their number of sex partners. Table 1 presents the determinants for this endpoint by different endpoint definitions.

Among socio-demographic determinants, MSM who were single but dating (adjusted odds ratio (aOR)=1.97, 95%CI=0.90;4.28) and who had an open/polyamorous relationship (aOR=1.86, 0.93;3.72) were more likely to be “probably/definitely intending” to reduce their number of sex partners. For the single endpoint, “definitely intending” to reduce the number of sexual partners, only MSM who were single but dating (aOR=0.06, 0.01;0.48) were less likely to reduce their number of sex partners. No behavioural determinants were found to be statistically associated. Among psycho-social determinants, for the combined endpoint MSM who knew anybody who has/had monkeypox (aOR=0.46; 0.27;0.80), who were more concerned about monkeypox infection (aOR=0.63; 0.53;0.77), who had a higher perceived risk of being infected by monkeypox (aOR=0.83; 0.68;1.02) and who had a more perceived problematic consequence of monkeypox (aOR=0.80, 0.64;1.00) were found to have lower odds of probably/definitely having fewer partner. However, these psycho-social associations were reversed (aOR=1.25 (0.54;2.88), aOR=2.54 (1.63;3,96), aOR=1.43 (1.01;2.02), and aOR=1.55 (1.00;2.39) respectively) when using the intention to definitely have fewer partners as the endpoint.

### Intention and determinants of the reduction of sexual activity

For the endpoint intention to have less sex, 309 (78%) MSM showed probable/definite intentions, and only 21 (5%) MSM showed a definite intention to reduce their number of sex partners. Table 2 presents the determinants for this endpoint by different endpoint definitions.

Among socio-demographic and behavioural determinants, MSM who were single but dating aOR=2.45, 95%CI=0.99;3.41), who had a monogamous relationship (aOR=2.83, 0.93;8.59), who had an open/polyamorous relationship (aOR=5.49, 2.56;11.76;8.59), who had 2-6 (aOR=10.21, 1.51;68.98), 7-15 (aOR=5.69; 0.81;40.17) and more than 15 sex partners (aOR=7.15; 0.94;54.51) were more likely to be probably/definitely intending to reduce their sexual activity. No such associations were obtained for the single endpoint, definitely intending to reduce sexual activity. Furthermore, neither socio-demographic nor behavioural determinants were found to be statistically associated. Among psycho-social determinants, only MSM who were more concerned about a monkeypox infection were associated with both probable/definite less sex intention (aOR=0.67, 0.54;0.83) and definite less sex intention (aOR=2.24, 1.36;3.70). again, the there was a different association obtained per endpoint definition, similar to the fewer partner intention presented above.

What is noteworthy to mention is that, for both endpoints, although MSM who had never used PrEP (PrEP naïve) or MSM who discontinued using PrEP (PrEP discontinued) were more likely to reduce their number of sex partners univariably, this association disappeared in the multivariable models.

## Discussion

Our findings based on 394 MSM in the Netherlands showed that 69% of our respondents had the intention to reduce their number of sexual partners probably or definitely, and 78% had the intention to probably or definitely have less sex in the context of the monkeypox epidemic.

While these descriptive figures look promising at first glance, the group of participants with definite intentions was remarkably smaller. Also, a closer inspection of the determinants reveals a more complex picture. Not surprisingly, dating or open relationship status plays a role in the intention to reduce the number of partners and sexual activity. Unexpectedly, no behavioural determinant tapping into past sexual behaviour or substance use turned out significant. For both endpoints, reduction of sex partners and having less sex overall an interesting distinction was observed. Definite intentions to reduce partners and sexual activity were positively influenced by psycho-social determinants, especially the concern to get infected by monkeypox, probable and definite intention jointly together as an endpoint show the reverse trend. Put differently, the two scale points, “probably yes” and “definitely yes” appear to be not on a continuum expressing intentions but are most likely qualitatively different statements with reversed (or large differences in magnitude of) determinants: Higher levels of infection concerns lead to weaker intention expressions to reduce the number of sexual partners and sexual activity, and only for those participants fully determined to have fewer sexual partners and have less sex there was a positive, facilitating relationship found. Probable intentions can be associated with underlying doubts and these doubts lead to the postponement of decisions to have less sex (partners).

Furthermore, it is noteworthy that PrEP naïve MSM and those that had discontinued with PrEP showed a higher (univariate) likelihood to reduce both the number of sex partners and sexual activity. This finding underlines the need to vaccinate the highest at-risk groups as they may not change their behaviour and at the same time inform PrEP users about the need to also display behaviour change, especially when they are not yet vaccinated against monkeypox.

## Conclusions

Based on our findings we conclude that the imposed demands by WHO to change one’s sexual behaviour at least among MSM in the Netherlands may not prove to be as easy and successful as necessary. Additional research is indicated to fully unravel underlying doubts associated with those behaviour change demands. Public health campaigns should have more emphasis on the necessity for this temporary behaviour change, to in turn target the population better. Those showing considerable levels of concern about a monkeypox infection may need to be exposed to additional arguments that would convince them to reduce their number of sex partners and to reduce sexual intercourse should they not be fully convinced. Likewise, unvaccinated PrEP users may need to understand that the sexual freedoms they enjoy in the context of HIV risk, may put them at considerable risk of an monkeypox infection.

In sum, to be able to curb the monkeypox epidemic successfully, both behavioural change (reduction of sex partners and less sex) and bio-medical approaches (vaccinations) are necessary. Yet achieving both endpoints requires public health messages and targeted campaigns that take note of the complexity of persuasive processes at play.

## Data Availability

Data are available upon request.

## Reference

1. Bragazzi NL, Kong JD, Mahroum N, Tsigalou C, Khamisy-Farah R, Converti M, et al. Epidemiological trends and clinical features of the ongoing monkeypox epidemic: A preliminary pooled data analysis and literature review. J Med Virol. 2022.

2. Bunge EM, Hoet B, Chen L, Lienert F, Weidenthaler H, Baer LR, et al. The changing epidemiology of human monkeypox—A potential threat? A systematic review. PLOS Neglected Tropical Diseases. 2022;16(2):e0010141.

3. ECDC. Joint ECDC-WHO Regional Office for Europe Monkeypox Surveillance Bulleten 2022 [Available from: https://monkeypoxreport.ecdc.europa.eu/.

4. RIVM. Monkeypox 2022 [Available from: https://www.rivm.nl/en/monkeypox.

5. RIVM. Monkeypox vaccination 2022 [Available from: https://www.rivm.nl/en/monkeypox/vaccination.

6. Kupferschmidt K. Monkeypox vaccination plans take shape amid questions. Science. 2022;376(6598):1142–3.

7. Miura F, van Ewijk CE, Backer JA, Xiridou M, Franz E, Op de Coul E, et al. Estimated incubation period for monkeypox cases confirmed in the Netherlands, May 2022. Euro Surveill. 2022;27(24).

8. WHO. Responding to the monkeypox outbreak: perspectives of clinicians treating patients with the disease. 2022.

9. CNN. WHO chief advises men who have sex with men to reduce partners to limit exposure to monkeypox 2022 [Available from: https://edition.cnn.com/2022/07/27/health/who-monkeypox-msm-sex-partners/index.html.

10. GGD-Amsterdam. Amsterdam start maandag 25 juli met vaccineren tegen monkeypox 2022 [Available from: https://www.ggd.amsterdam.nl/nieuwsoverzicht/start-vaccineren-monkeypox/.

11. Wang H, Abreu de Paulo KJI, Gultzow T, Zimmermann HML, Jonas KJ. Monkeypox self-diagnosis abilities, determinants of vaccination intention and self-isolation intention after diagnosis among MSM in the Netherlands. medRxiv. 2022:2022.07.29.22278167.

